# Clinical actionability of genetic findings in cerebral palsy

**DOI:** 10.1101/2023.09.08.23295195

**Authors:** Sara A Lewis, Maya Chopra, Julie S. Cohen, Jennifer Bain, Bhooma Aravamuthan, Jason B Carmel, Michael C Fahey, Reeval Segel, Richard F Wintle, Michael Zech, Halie May, Nahla Haque, Darcy Fehlings, Siddharth Srivastava, Michael C Kruer

## Abstract

**Background and objectives:** Single gene mutations are increasingly recognized as causes of cerebral palsy (CP) phenotypes, yet there is currently no standardized framework for measuring their clinical impact. We evaluated Pathogenic/Likely Pathogenic (P/LP) variants identified in individuals with CP to determine how frequently genetic testing results would prompt changes in care.

**Methods:** We analyzed published P/LP variants in OMIM genes identified in clinical (n = 1,345 individuals) or research (n = 496) cohorts using exome sequencing of CP patients. We established a working group of clinical and research geneticists, developmental pediatricians, genetic counselors, and neurologists and performed a systematic review of existing literature for evidence of clinical management approaches linked to genetic disorders. Scoring rubrics were adapted, and a modified Delphi approach was used to build consensus and establish the anticipated impact on patient care. Overall *clinical utility* was calculated from metrics assessing *outcome severity* if left untreated, *safety/practicality* of the intervention, and anticipated intervention *efficacy*.

**Results:** We found 140/1,841 (8%) of individuals in published CP cohorts had a genetic diagnosis classified as *actionable*, defined as prompting a change in clinical management based on knowledge related to the genetic etiology. 58/243 genes with P/LP variants were classified as actionable; 16 had treatment options targeting the *primary disease mechanism*, 16 had *specific prevention strategies*, and 26 had *specific symptom management* recommendations. The level of evidence was also graded according to ClinGen criteria; 44.6% of interventions had evidence class “D” or below. The potential interventions have *clinical utility* with 97% of outcomes being moderate-high *severity* if left untreated and 62% of interventions predicted to be of moderate-high *efficacy*. Most interventions (71%) were considered moderate-high *safety/practicality*.

**Discussion:** Our findings indicate that actionable genetic findings occur in 8% of individuals referred for genetic testing with CP. Evaluation of potential *efficacy*, outcome *severity*, and intervention *safety*/*practicality* indicates moderate-high *clinical utility* of these genetic findings. Thus, genetic sequencing to identify these individuals for precision medicine interventions could improve outcomes and provide clinical benefit to individuals with CP. The relatively limited evidence base for most interventions underscores the need for additional research.

## INTRODUCTION

A genetic etiology for 15-34% of cerebral palsy (CP) cases can be identified using exome sequencing (ES) ^1^ and commercial CP gene sequencing panels are available for clinical use. Although clinicians are increasingly using both panels and ES as part of the diagnostic workup of CP patients, the impact of genetic findings on clinical care is still being determined. The current genomic landscape of CP is broad, with hundreds of individually rare single-gene or Mendelian causes. ^1^. Given the increased utilization of clinical genetic testing ^2^ and the heterogeneity of CP-associated genes being identified, there is a need to evaluate how genetic findings can impact care for individuals with CP.

Prior studies have assessed the actionability of genetic findings in epilepsy using an iterative process of expert review and curation. A recent study of 2,008 adults with epilepsy reported that 11% of their cohort had identifiable causative mutations. In 56% of these cases, the genetic finding was expected to be actionable, prompting a change in clinical management^3^. In addition, one downstream benefit of identifying potentially actionable genetic findings is the development of targeted gene sequencing panels to identify individuals who could benefit from specific treatments.

ClinGen (www.clinicalgenome.org) is a resource supported by the U.S. National Institutes of Health. Its goal is to define the clinical relevance of genes and their variants for use in precision medicine and research. A ClinGen working group previously published a landmark rubric to assess the clinical actionability of genetic results ^4^. However, this rubric focused on assessing incidental/secondary findings identified during clinical genetic sequencing rather than etiologic findings identified during a diagnostic workup.

By analyzing published data, we sought to build on prior efforts to determine to what extent genetic findings could impact the clinical care of individuals with CP. A working group comprised of content experts was organized and began with the established framework for determining actionability employed by studies of epilepsy genetics. A systematic review of the literature and synthesis of evidence was used to compile a list of genes with actionable genetic findings. We then incorporated the ClinGen rubric and modified it to apply to the diagnostic workup of a person with CP in a pediatric care setting. We recognized that additional measures of clinical impact would provide added value for practicing clinicians. Accordingly, we developed estimates of untreated *outcome severity, safety/practicality* (of a particular intervention), and anticipated intervention *efficacy*. We then calculated an overall *clinical utility* score by combining these measures. Our overall objective was to determine the potential impact of identifying a genetic etiology on clinical management in CP. Our findings, including curated lists of genes, outcomes, and available interventions, form a starting point for using genetic data and supports the utility of this testing for improved outcomes for individuals with CP.

## METHODS

### Cohorts and variant calling

An overview of the study methodology is described in **Figure 1**. We identified four published research cohorts ^5-8^ (aggregate n = 496 trios) meeting inclusion criteria of at least 10 individuals with a cerebral palsy diagnosis receiving trio-based ES and variant pathogenicity classified according to ACMG guidelines ^9^. We also included a clinical cohort referred to GeneDx for ES with suspected or confirmed CP, including 1,009 individuals analyzed as trios and 336 as singletons ^10^. For pooled analysis, CP-associated genes from the research cohort were filtered to match the gene list described in Moreno-De-Luca et al ^10^.

**Figure 1:**
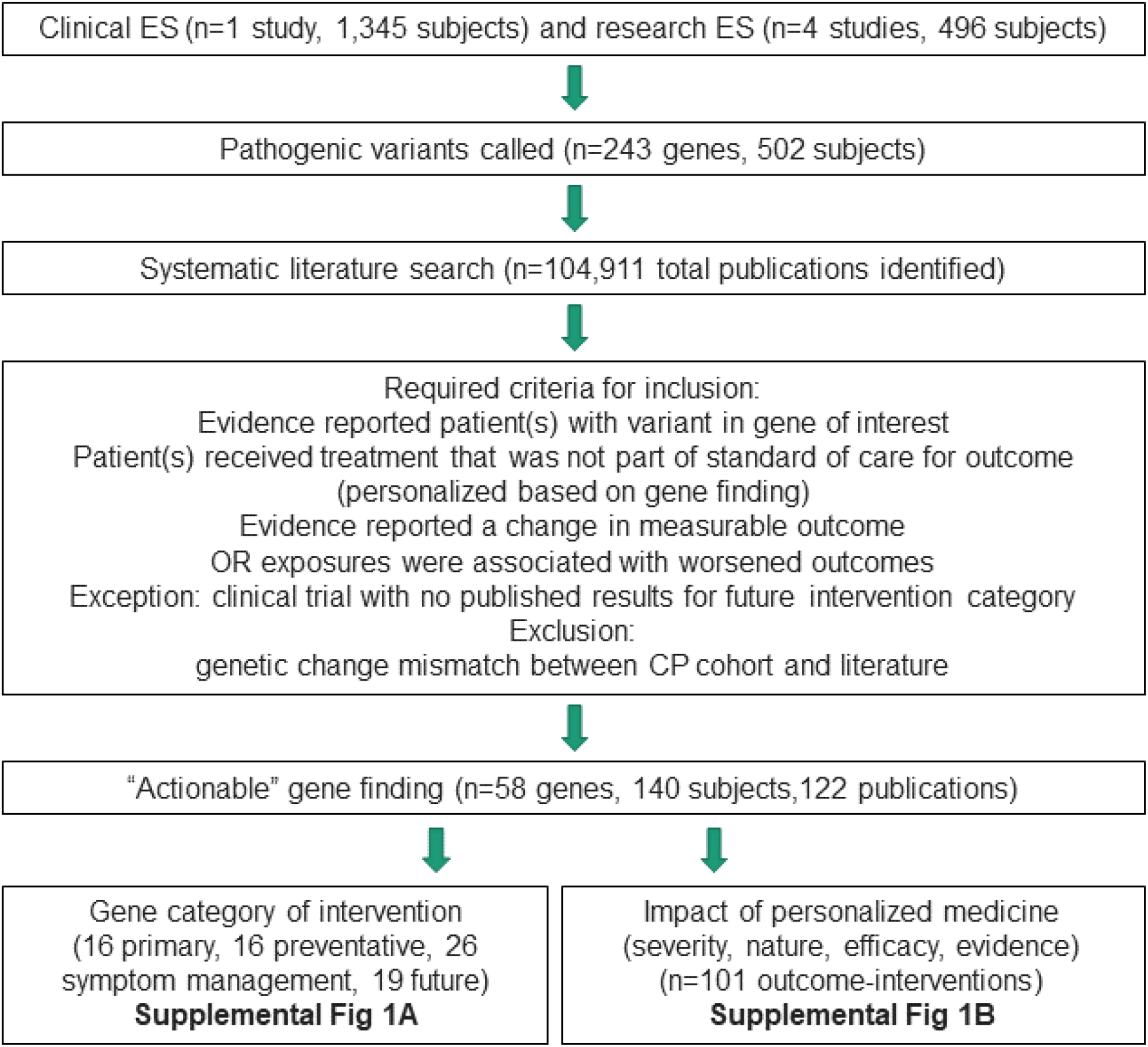
Actionable cerebral palsy gene literature search strategy and workflow. See Supplemental Methods for query terms and tools used for literature review. Publications were eliminated if patients did not have variants in listed gene or if interventions were not trialed in patients directly, if treatment was part of standard of care, if potential outcome had no effect on patient function, or if the reported phenotypes were not consistent with the genetic disorder in that gene most closely aligned to CP. Most of the identified literature (n=104,789 publications) did not meet inclusion criteria for a personalized medicine intervention based on genetic finding. Category assignment criteria and process described in **Supplemental Figure 1A**. Clinical utility impact assessment criteria and process described in **Supplemental Figure 1B**.

### Systematic literature review and inclusion criteria

PRISMA standards were used to guide literature review^11^. Publicly available tools (clinicaltrials.gov, Drug-Gene Interaction Database, GeneReviews, GeneRx, Mastermind, PubMed) were queried between August 2021-July 2022 to identify treatments or interventions that could be considered based on genetic findings. Category definitions, search parameters, and links to web-based databases and questionnaires are available in **Supplemental Materials**. Publications were required to be peer-reviewed, include patients with variants in the gene of interest, and test an intervention that led to a noticeable improvement in a defined outcome. Irrelevant instances (i.e., literature where the identified genetic disorder was was unrelated to neurodevelopment) were excluded.

### Actionability definitions and categories

We adapted the ClinGen protocol for evaluating available evidence and utilizing a semiquantitative metric to describe the actionability of genetic findings ^4^. We defined a genetic finding as *actionable* if it was able to be classified as having an intervention that fell into at least one of the three categories defined as follows:

#### Primary disease mechanism

intervention directly affecting the gene product, a supplement for a primary deficiency, or a specific treatment for a primary cause of pathology.

#### Specific prevention strategies

known, specific triggers associated with the genetic abnormality that worsen the condition and could be decreased through avoidance, early detection, or lifestyle changes.

#### Specific symptom management

evidence that a medication or intervention improves symptoms associated with the disorder.

A gene may have interventions in different categories but was assigned to a single *actionability* category in rank order: *primary disease mechanism, specific prevention strategy, or specific symptom management. Potential for future intervention* (an intervention undergoing a clinical trial listed in clinicaltrials.gov or included in the Drug-Gene Interaction database) was selected only if the intervention in question did not meet the criteria for another category.

Clinical trials had to be interventional in nature and in either Phase II or III. Such instances were not considered in the overall calculation of *actionability* but were reported as a separate category. In the future, such interventions could be re-classified in one of the three actionability criteria. Other classes of actionability, such as family planning or referrals to different specialists, were not considered.

### Defining outcomes/interventions and scoring clinical utility

101 outcome/intervention pairs were curated using the abovementioned tools and strategies. An outcome is a potential or ongoing clinical concern affecting the patient. An intervention was defined as “actionable” when knowledge of a genetic etiology would be anticipated to prompt a change from standard care. Standard care was defined here as the typical treatment for the clinical manifestation/outcome regardless of etiology. For disorders where many drugs are available (i.e., antiepileptics), reports of efficacy for a specific drug were considered actionable as precision medicine approaches could include the selection of a specific medication rather than adopting a ‘trial-and-error’ approach. A list of definitions is provided in **Supplemental Table 1**.

*Clinical utility* metrics were derived from the ClinGen rubric ^4, 12^ and adjusted for relevance in a pediatric neurology/neurodevelopmental care setting using the following grounding scenario:

> Imagine you are a clinician treating a young child diagnosed with CP. If an actionable genetic finding was identified, how could information about the seriousness of the condition’s natural history (*outcome severity*), risk and/or burden on the patient/family associated with the actionable intervention (*safety/practicality)*, or potential benefit of the actionable intervention *(efficacy)* impact your decision-making?

*Clinical utility* scores were individually generated, and the strength of available evidence was considered a separate category independently of other factors. The relative frequencies of outcomes or adverse events were not explicitly factored into scoring, as we could not determine whether a given outcome was present in individuals from this aggregate cohort. *Clinical utility* scores were then defined as follows:

#### Outcome severity

impact of the condition on patient function if left untreated.

#### Safety/Practicality

impact of a potential adverse event (AE) and/or disruption to patient/family, such as invasiveness, need for lifestyle alterations, or specialized monitoring or management.

#### Efficacy

degree to which an intervention can prevent or reduce symptoms, improve function, or decrease the likelihood of additional complications/manifestations.

#### Clinical utility

an aggregate score calculated by adding *outcome severity, safety/practicality*, and *efficacy* scores. Each metric used a 0-3 scale, with higher scores corresponding to more compelling reasons for clinical intervention.

We developed guidelines for scoring clinical utility categories using modifications of the ClinGen rubrics (**Table 3**). The Common Terminology Criteria for Adverse Events (CTCAE v5.0) (https://ctep.cancer.gov/protocoldevelopment/electronic_applications/docs/ctcae_v5_quick_reference_5×7.pdf) from the U.S. Department of Health and Human Services and the Modified Treatment Burden Questionnaire (MTBQ) were incorporated into the *safety*/*practicality* subscale. Draft rubrics were circulated for group feedback and finalized using a modified Delphi process. Grounding examples were discussed during the development of the rubric, and examples are given in **Table 3**. We solicited individual scores and used a threshold of 3 or more scorers in agreement as indicative of consensus. Scorers were permitted to skip entries, and additional scorers were recruited when more scores were needed to achieve consensus; therefore, each score could be assessed by anywhere from 3-9 scorers. In instances without an agreement, the scoring was iteratively discussed as a group, and modifications were made to the rubric until consensus was reached. 3 rounds were required for severity, 2 rounds for safety/practicality, and 2 for efficacy.

Graphs were created using the tidyverse library with jitterplot function in R (4.2.2). For box and whisker plots, box indicates 75th and 25th percentile with 50% median line; whiskers indicate range of data. Statistical tests including chi square and 2 tailed t-tests for equal variance were also calculated in R. Inter-rater reliability was calculated using the Gwet AC2 coefficient, percent agreement, and statistical significance ^13^ using the irrCAC library in R (4.2.2). Percent agreement was defined as the number of scores in agreement compared to the total scores for that metric (tolerance=0). The AC2 statistic is corrected for potential agreement by chance as well as agreement with classification errors. Inter-rater reliability and percent agreement calculations were conducted independently on subsets based on the metric scored and the number of raters.

### Standard Protocol Approvals, Registrations, and Patient Consents

Study was reviewed by the Phoenix Children’s Hospital IRB (IRB-23-260) and determined to be exempt from human subject research oversight and granted a waiver for patient consent.

### Data availability

Data generated during this study including raw scores and Pubmed sampling script available on GitHub https://github.com/Kruer-Lab/actionability with access provided upon request.

## RESULTS

### Clinical actionability

Our aggregate cohort included 1,841 individuals, for whom pathogenic or likely pathogenic (P/LP) variants were identified in 502, for an overall diagnostic yield of 27%. These variants were distributed among 243 unique genes (**Supplemental Table 1**); 94 genes were recurrent, with ≥2 affected individuals that met ACMG criteria for P/LP variants consistent with the known inheritance model for each gene.

We found that 58/243 (24%) of genes with P/LP variants in our CP cohort could be classified as *actionable* (**Table 1**). Another 16/243 (7%) of genes were considered to have *the potential for future intervention*. 140/1,841 (8%) of all individuals who underwent ES had a genetic finding expected to alter their management. Thirteen additional individuals had a genetic result with treatments in clinical trials, and a further 18 had gene findings represented in a preclinical development database. Together, 171/1,841 (9%) of individuals with CP who underwent ES had a genetic finding with precision medicine treatments either currently available or in development (**Fig. 2A**).

**Table 1.** Frequency of actionable genes and patients with genetic findings by categories of interventions. Likely pathogenic or pathogenic variants (P/LP) called by authors using in-house methods and ACMG guidelines. Percentage of patients with a treatable variant in that class determined by dividing by the number of patients in that group with a P/LP variant. 58/243 genes (24%) are considered treatable and 140/502 (28%) of patients with P/LP findings had currently available treatments (defined as primary, specific symptom, or preventative).

|  | **Primary disorder** | **Specific preventative strategies** | **Specific symptom management** | **Future (Clinical Trials or Druggable Genome)** |
| --- | --- | --- | --- | --- |
| Total Genes | 16/243 (6.6%) | 16/243 (6.6%) | 26/243 (10.7%) | 19/243 (7.8%) |
| Clinical (%) | 15/224 (6.7%) | 16/224 (7.1%) | 24/224 (10.7%) | 18/224 (8.0%) |
| Research (%) | 2/50 (4.0%) | 3/50 (6.0%) | 6/50 (12.0%) | 5/50 (10.0%) |
| Total Patients | 31/502 (6.2%) | 33/502 (6.6%) | 76/502 (15.1%) | 38/502 (7.6%) |
| Clinical Patients | 28/431 (6.5%) | 29/431 (6.7%) | 66/431 (15.3%) | 33/431 (7.7%) |
| Research Patients | 3/71 (4.2%) | 4/71 (5.6%) | 10/71 (14.1%) | 5/71 (7.0%) |

**Figure 2.**
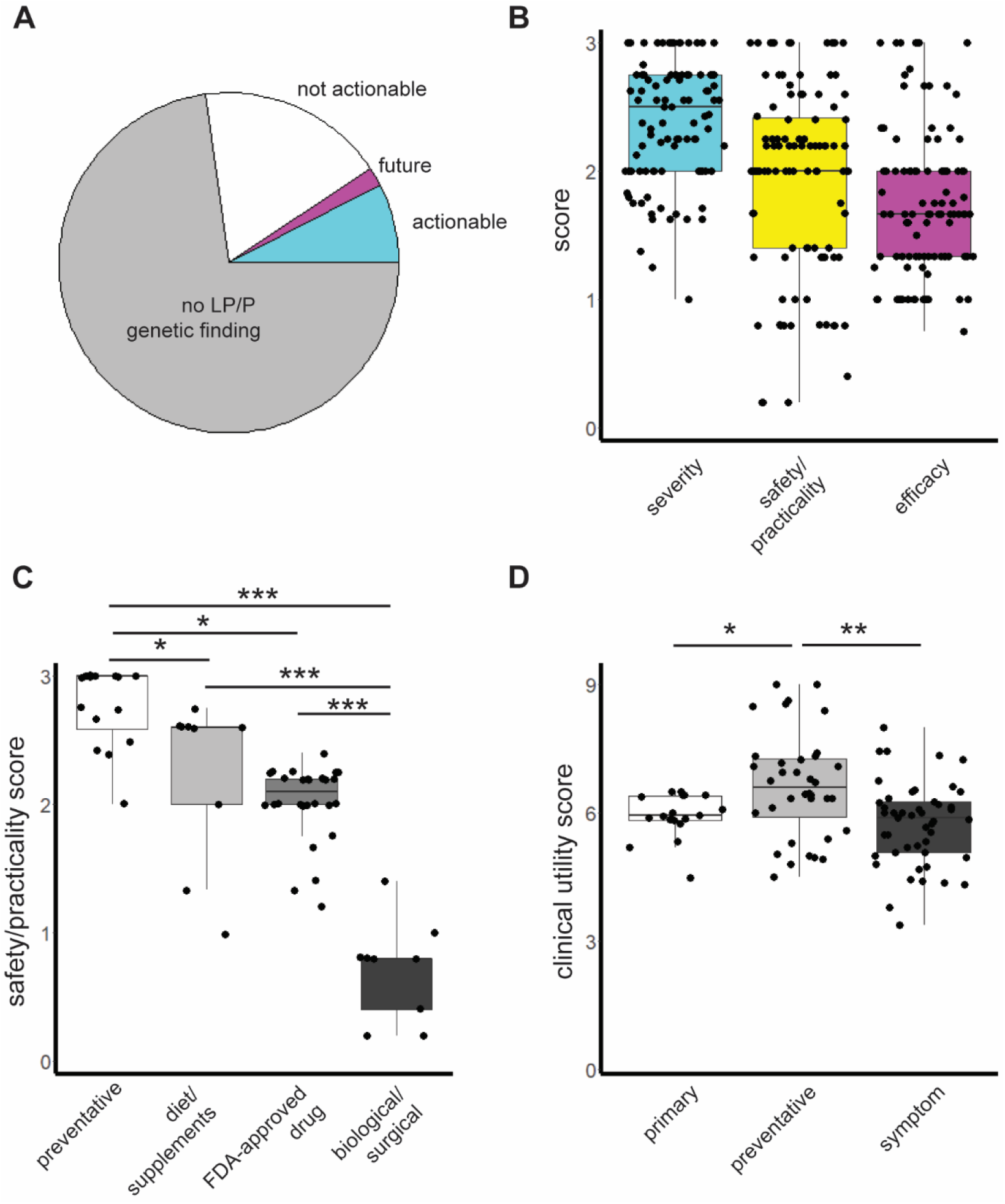
Frequency and scores for actionable findings. **A**, Percentage of cohort with P/LP variant calls and with current for future actionable gene findings identified via literature review. **B**, Scores for clinical utility metrics of severity, safety/practicality, and efficacy averaged across all scorers. **C**, Scores for safety/practicality averaged across all scorers for 61 unique interventions by category including preventative, dietary changes or adding over-the-counter supplements, FDA-approved drugs, and biological/surgical interventions including gene or cellular therapy, enzyme replacement therapy, or surgical interventions. **D**, Total clinical utility score for each category of intervention. Preventative interventions had a higher clinical utility than interventions targeting the primary disease mechanism or specific symptom management. Box represents 25 and 75 percentile, the horizontal line represents the median, and whiskers the range of data. n=101 total outcome-intervention pairs, 18 primary, 36 preventative, 47 symptom management interventions. * p=0.02, ** p=0.0003, ***<2.0×10^−5^ by 2 tailed t-test.

We found that the proportion of actionable findings was similar for research and clinical cohorts. There was no difference in the ratios of actionable genes (11/50 vs 55/224, p=0.66) or individuals with actionable findings (17/71 vs 123/431, p=0.98, both n.s. by chi-square) between these groups. These findings support the pooling of these datasets for our subsequent analyses.

### Clinical utility

We subsequently scored 101 outcome-intervention pairs for 58 distinct genes for their *clinical utility*. Our working group assessed *clinical utility* in the context of disease *outcome severity*, intervention *safety/practicality*, and intervention *efficacy*. A complete list of outcome-intervention pairs and assigned scores can be found in the **Supplemental Excel**, with average scores in **Figure 2** and summarized data for each category in **Table 2**.

**Table 2.** Summary scores of clinical utility categories. Numbers indicate n=101 total outcome/interventions.

|  | **Severity** | **Safety/Practicality** | **Efficacy** | **Combined** |
| --- | --- | --- | --- | --- |
| **0 (no impact)** | - | 4 | 0 |  |
| **1** | 3 | 25 | 38 |  |
| **2** | 43 | 48 | 45 |  |
| **3 (most impact)** | 55 | 24 | 18 |  |
| **Average** | 2.40 | 1.94 | 1.76 | 6.10 |
| **CI** | 2.30-2.49 | 1.80-2.08 | 1.65-1.87 | 5.89-6.31 |

#### Outcome Severity

We considered the natural history associated with OMIM clinical phenotypes that best matched the disease mechanism for each gene and scored outcomes as mild, moderate, or severe based on rubric criteria (**Table 3**). We considered short and long-term scenarios to capture day-to-day functional impairments and the potential for worsening function, episodic decompensations, or the possibility of premature death. We only considered outcomes that had evidence of being modified by the intervention. We identified 55 severe outcomes, 43 moderate outcomes, and 3 mild outcomes (**Fig 2B**). The average *outcome severity* score was 2.40 (95 CI 2.30-2.49, **Table 2**) on our scale of 0 to 3. This suggests that outcomes associated with variants in these genes may substantially impair function, supporting the potential benefit of identifying disease-modifying interventions.

#### Safety/Practicality

We assessed the safety and/or practicality of potential interventions (**Table 3**). We evaluated *safety* as the risk posed by possible adverse events (AE) and *practicality* as the degree of burden of the intervention. We evaluated safety based on the type of interventions. We found that preventative measures, dietary changes, and FDA-approved compounds generally scored well. In contrast, experimental interventions or those requiring special monitoring, such as gene-replacement and surgical therapies, generally scored poorly (**Fig 2C**). We identified 24 no- or low-risk, 48 mild-risk, 25 moderate-risk, and 4 high-risk interventions (**Fig 2B**). Overall, many available treatments feature relatively acceptable risk profiles and moderate burden for individuals, with an average score of 1.94 (95% CI 1.80-2.08, **Table 2**).

#### Efficacy

We then assessed the ability of a successful intervention to reduce symptoms or avoid an unwanted outcome (**Table 3**). This score describes the relative impact of a successful prevention or treatment. It could be based on a quantitative measurement, such as a clinical trial outcome, or a qualitative statement by the author describing the change to the patient outcome. Intervention efficacy was not weighted by the strength of the evidence supporting its use (see below). We identified 18 highly effective, 45 moderately effective, and 38 mildly effective interventions (**Fig 2B**). The overall efficacy was 1.76 (95% CI 1.65-1.87, **Table 2**) suggesting that these interventions can modify the severity of the potential outcome.

#### *Clinical utility* overall score

We found an average score of 6.10 (95% CI 5.89-6.31, **Table 2**). The range was 3.38-9.0 from a possible range of 0-9. We compared clinical utility scores between different categories of interventions and found that preventative interventions were associated with higher clinical utility scores than targets of the primary mechanism of disease (6.65 vs. 5.94, p=0.02; t-test) and symptomatic management strategies (6.65 vs. 5.75, p=0.0003; t-test) (**Fig 2D**). There was no difference in clinical utility for genetic findings with primary disease targeting interventions and symptom management (5.94 vs 5.57 p=0.48; t-test). This suggests that the overall clinical utility of identifying a genetic etiology and implementing precision medicine strategies is moderate to high for CP.

#### Inter-rater reliability

We calculated the percent agreement between scorers for severity, safety/practicality, and efficacy, based on the number of scorers contributing to the average score (**Supplemental Table 3**). We identified an average of 85.5% agreement (95% CI 81.0-89.6%). When we assessed the inter-rater reliability for each metric using Gwet AC2, we found an AC2 average rating of 0.66 (95% CI 0.56-0.77), which is considered to be substantial agreement (between 0.60-0.79). We also calculated the weighted average for AC2 based on the number of outcomes or interventions and determined it to be 0.75. This agrees with the observation that lower agreement scores corresponded to a subset of items requiring multiple scoring rounds, consensus discussions, and adding more scorers. All comparisons had statistically significant agreement, except for 2 efficacy outcome-interventions that required 6 scorers to achieve consensus.These findings demonstrate substantial agreement between scorers when scoring the potential outcome severity, interventional risk/burden, and intervention efficacy using the designed rubrics.

### Levels of evidence

Evidence was weighted on an alphabetical ranking using modified ClinGen criteria drawn from Oxford Centre for Evidence-Based Medicine guidelines (**Table 4**). Notably, 56/101 (55.4%) of outcome-intervention pairs had a C or greater level of evidence supporting a given prevention or treatment.

**Table 3:** Rubric for scoring impact of genetic finding. Severity evaluates disease-related outcomes for 1) short-term impact based on outcome effect on comfort, development, and/or adaptive function 2) long-term impact based on potential functional decline, decompensations, or risk of serious events. The single most appropriate severity category was assigned for each outcome. Outcomes that had no impact on function (0 on the 0-3 scale) were not considered in this study. Risk assesses severity of a potential adverse event (AE) as it is not possible to reliably ascertain absolute risk due to missing frequency data. Risk classifications also use Common Terminology Criteria for Adverse Events (CTCAE) v5.0 from U.S. Department of Health and Human Services. Practicality considers examples from the Multimorbidity Treatment Burden Questionnaire (MTBQ) to assess impact of treatment on patient’s lifestyle. ADL=activities of daily living. AE=adverse events. Efficacy evaluates the extent to which an intervention can prevent or significantly reduce symptoms, improve function, or decrease risk of additional complications/manifestations. Interventions with no efficacy (0 on the 0-3 scale) were not considered in this study.

| **Metric description** | **Score and definition** | **Example** |
| --- | --- | --- |
| Severity: short-term impact (active clinical management) | 1=Impairments present, but do not substantially impact adaptive function or self-help skills (mild) | Joint stress |
|  | 2=Impairments limit adaptive function and self-help skills (moderate) | *NKX2-1*-associated chorea |
|  | 3=Complete lack of adaptive function and self-help skills (severe) | *PANK2*-associated dystonia |
| Severity: long term impact (outcome or risk for severe events if untreated) | 1=Transient decompensations (mild) | Paroxysmal dyskinesia |
|  | 2= Decompensations or manifestations can worsen function, but may be small changes or not permanent (moderate) | *ATP1A3*-associated episodic decompensation |
|  | 3= Decompensations or manifestations can permanently worsen function or are a serious event (severe) | Sudden cardiac death |
| Safety: Potential risk of intervention | 0= Life-threatening consequences; urgent intervention indicated OR Death related to AE (CTCAE Grade 4-5) | Stem cell transplantation (organ failure, sepsis) |
|  | 1= Severe or medically significant but not immediately life-threatening; hospitalization or prolongation of hospitalization indicated; disabling; limiting self-care ADL (CTCAE Grade 3) | DBS surgery (peri-lesional edema, mild intracerebral hemorrhage, infection) |
|  | 2= Moderate; minimal, local or noninvasive intervention indicated; limiting age-appropriate instrumental ADL (CTCAE Grade 2) | Levodopa (vomiting) |
|  | 3 = Mild; asymptomatic or mild symptoms; clinical or diagnostic observations only; intervention not indicated (CTCAE Grade 1) | leucine supplements (clinically-silent hypoglycemia) |
| Practicality: The impact of intervention on patient/family | 0 = Highly invasive |  |
|  | 1 = Invasive, significant impact on lifestyle, or specialized management required | Low valine/protein diet |
|  | 2 = Potential for burden, but reasonable for most patients; treatment does not require intensive oversight | Ketogenic diet |
|  | 3 = Minimum burden and/or oversight | Avoid contact sports |
| Efficacy: Improvement of outcome with intervention | 1 =Small improvement in condition or decrease in risk of acquiring potential outcome | Levodopa for *CTNNB1*-associated dystonia |
|  | 2 = Moderate improvement in condition or decreased risk in acquiring potential outcome | Caffeine for *ADCY5*-associated dyskinesia |
|  | 3 = Significant improvements, complete avoidance of potential outcome | Levodopa for *SRP*-associated dopa responsive dystonia; avoid metronidazole to prevent *ERRC6*-related hepatic failure |

**Table 4:** Grade of evidence. Definitions and grades reflect ClinGen criteria drawn from Oxford Centre for Evidence-Based Medicine guidelines.

| Grade | Definition | # of interventions |
| --- | --- | --- |
| **A** | Clinical guideline consensus statement/FDA approved for application | 19 |
| **B** | Literature review with multiple case-control studies | 36 |
| **C** | Case-control multi-patient trial with quantitative evidence for outcomes | 1 |
| **D** | Multiple-patient clinical report or expert opinion without further information | 37 |
| **E** | Single-patient clinical report | 5 |
| **N** | Source not found | 3 |

## DISCUSSION

We found that 8% of the individuals within our CP cohort harbored a clinically *actionable* genetic finding. This is comparable to the 6% identification rate of clinically actionable findings reported in individuals with epilepsy (121/2008) ^3^. This has implications for the power of genetic testing in this patient population to identify personalized medicine approaches and improve patient outcomes.

In addition to the methodology used to assess the actionability of genetic findings in epilepsy, we also developed tools to evaluate the *clinical utility* of actionable interventions that could be employed in future studies. With these tools, we were able to quantitatively assess *clinical utility* to help further capture the impact that personalized medicine strategies could have on ongoing clinical care. The average *outcome severity* score was 2.40, representing a moderate-severe rating. Most interventions were judged to be tolerable for *safety/practicality*, with an average score of 1.94. Finally, the average *efficacy* score was 1.76. These results suggest that the interventions considered target relevant aspects of the disorder, are reasonably safe and acceptable, and have the potential for clinically significant impact. Preventative strategies yielded higher safety and clinical utility scores, suggesting that early identification of genetic etiologies will be necessary for optimal outcomes. We identified substantial inter-rater reliability with 85% agreement, suggesting that individuals with different training and specialties can utilize the developed rubrics to achieve similar clincal utility scores when used in parallel with consensus discussions.

Despite the potential *clinical utility* of *actionable* findings, most interventions are typically off-label. Although this is not unusual in pediatric neurology practice ^14^, it is notable that the evidence base for half (44.6) of the *actionable* interventions is limited, usually derived from small cohort studies, defined as class D or lower (**Table 4**). This highlights a need for additional studies of potential precision medicine approaches in CP using rigorous and reproducible designs. This is particularly important given that additional CP-associated genes are being discovered rapidly ^1, 15^ and that the list of potentially actionable genes is expected to grow in parallel. N-of-1 treatment trials have recently been proposed ^16^ as one possible solution to the evidence gap for neurological disorders, particularly individually rare neurodevelopmental disorders ^17^.

There are several important limitations to this work. First, this approach likely missed potential interventions reported in the literature that did not meet our search criteria. We also did not consider other changes to clinical management, such as referral to specialists. Our findings are likely an underestimate of the number of individuals with cerebral palsy who would benefit from genetic testing. Second, clinical presentations can vary considerably from one individual to the next despite a common genetic etiology. This can affect the applicability of actionable findings for a given gene. In addition, the severity of clinical symptoms may also vary from patient to patient. We recognized this inherent challenge and sought to emphasize typical presentations and symptom severity whenever possible and to strike a balance when considering possibilities rather than focusing on extremes. We used a modified Delphi process, employing frequent consensus-building discussions to clarify points of contention or discrepancies in scoring. Accordingly, we did not conduct a risk-benefit analysis for each outcome-intervention pair but evaluated metrics of *clinical utility* instead.

We included variants in genes that may result in phenotypes that are not associated with cerebral palsy, including progressive or regressive phenotypes and non-neurodevelopmental features. Given that such cases could be incompatible with the international consensus definition of CP ^18^, doing so may invite criticism that such individuals “do not truly have CP.” Conversely, “atypical” clinical phenotypes resembling CP within a broader physical neurodisability umbrella are increasingly recognized in related genetic disorders, such as hereditary spastic paraplegia ^19^. Considering such factors, we have retained these cases in our analysis because they were identified in research cohorts clinically ascertained as having CP or a clinical cohort referred based on suspicion of CP. This reflects real-world clinical practice where incomplete histories, longitudinal follow-up, or other factors can prompt a reevaluation.

ES and/or genome sequencing has recently been recommended by the American College of Medical Genetics & Genomics as a first tier test for individuals with congenital anomalies, (global) developmental delay, and/or intellectual disability, based upon the Grading of Recommendations Assessment, Development and Evaluation (GRADE) evidence framework ^20^. In the future, the methods and classification scheme utilized in this work could be readily extrapolated to conduct a similar analysis. We anticipate actionability will improve over time as the degree of evidence improves with additional case studies and clinical trials for interventions based on genetic etiologies. Thus, this study provides both a framework and initial data to support such future efforts to guide providers caring for individuals with CP.

## Conclusions

There is an increasing emphasis on early diagnosis and intervention for cerebral palsy for optimal outcomes ^21^. Although standard of care is improving for CP interventions including preventative, occupational, physical and speech therapies ^22^, there is a need for precision medicine as well. Case studies, a review of some common genes with known effective interventions, and site-specific reports of changes in clinical management have been reported elsewhere^8, 23^. Here, we introduce a systematic approach to evaluating clinical utility for CP genetic findings, a list of resources, and estimated frequency of patients who would benefit. In our cohort, approximately one-third of the *actionable* interventions include preventative measures, suggesting that medical interventions may increasingly represent an important early intervention opportunity. Early utilization of genetic sequencing may thus lead to the best possible clinical outcomes, particularly when considering missed treatment opportunities that could ameliorate functional impairments and reduce lifetime care costs to the families, payers, and society, which can be considerable ^24^. Our findings indicate potential opportunities for tailored treatments in CP based on genetic findings and identify opportunities for precision medicine to improve outcomes in individuals with CP, laying the groundwork for additional, larger-scale studies.

## Data Availability

Data generated during this study including raw scores and Pubmed sampling script available on GitHub with access provided upon request.

https://github.com/Kruer-Lab/actionability

## ACKNOWLEDGEMENTS

The authors thank Tyler Kruer for custom Pubmed query scripts and GitHub repository assistance.

## AUTHOR CONTRIBUTIONS

Conceptualization: SAL, MCF, MCK

Design: SAL, MC, JSC, SS, DF, MCK

Individual scoring: MC, JSC, JB, BA, JBC, RS, MCF, DF, SS, MCK

Consensus scoring: SAL, RFW, HM

Literature review: SAL, HM, NH

Writing: SAL, MCK

Revisions: all

