## Supplemental materials for "Clinical actionability of genetic findings in cerebral palsy"

### SUPPLEMENTAL MATERIAL

#### Supplemental Methods

#### Search strategy to identify actionable findings

Tools were accessed independently by either HM, NH, or SAL. All potential evidence was assessed by review team of all authors to ensure it matched the inclusion/exclusion criteria and that the mechanism of disease and treatment aligned (below).

Clinical Genome Resource Actionability Knowledge Repository Center (https://actionability.clinicalgenome.org/ac/) Search parameter: gene name. Inclusion criteria: must relate to pediatric disorder. This resulted in 2 hits.

NIH GeneReviews books (https://www.ncbi.nlm.nih.gov/books/) search parameters: gene name. Inclusion requirement: Must list specific treatment rather than standard management for the condition. Last accessed March 2022. The number of books meeting the search criteria in were not tracked at the time of the initial review. Instead, an estimate was created August 2023 using a custom script with the terms “gene name” and “treatment”. This resulted in 6,208 books for 207 genes. This script is available at the Github link described in the data sharing statement.

GeneRx[24] (https://www.rx-genes.com/) search parameters: gene name. Last accessed October 2021. 30 entries.

Pubmed (https://pubmed.ncbi.nlm.nih.gov/) search parameters: [gene name or OMIM phenotype term] AND [treatment or intervention]. If needed, results are filtered by adding AND patient and/or AND [specific outcome]. Initial inclusion in actionable gene list search concluded March 2022. For some genes, additional evidence was identified during the efficacy review process by DF, JB, SS, SAL, or MCK (last accessed July 2023). The number of studies meeting the search criteria in Pubmed were not tracked at the time of the initial review. Instead, an estimate was created August 2023 using a custom script with the terms “gene name” AND (either) “treatment” (or) “intervention” AND “patient”. This resulted in 97,689 hits. The script is available at the Github link described in the data sharing statement.

Mastermind[25] (https://www.genomenon.com/mastermind/) search parameters: [gene name] AND [text=treatment] AND [disease=Cerebral palsy]. Filters: +treatment +therapy +prognosis. Last accessed February 2022 by HM. 1014 entries for 148 genes.

Clinical trials (Clinicaltrials.gov) search parameters: gene name AND disorder name(s). Inclusion criteria: phase II/III studies, interventional study design (compounds or treatments), and either enrolling or concluded studies. Exclusion criteria: terminated studies. Last accessed July 2023 by NH and SAL. 769 entries for 16 genes.

Drug Gene Interaction Database v3.0 [26] (http://www.dgidb.org/) search parameters: gene name Inclusion criteria: >1 interaction score. Exclusion criteria: drug has no citation/evidence. Last accessed August 2021. 27 entries for 9 genes.

**Mechanism of disease and treatment alignment process:** A combination of *de novo*, recessive, and X-linked pathogenic/likely pathogenic (P/LP) variants were identified in the four cohorts. Given this mixed inheritance pattern, we considered haploinsufficiency and/or loss of function the predominant molecular mechanisms represented at the cohort level for evaluating the evidence. We identified OMIM phenotype numbers for the 58 potentially actionable genes most closely related to CP for the outcomes of interest for conducting the literature search. We scored severity by aligning reported disease manifestations for the assigned OMIM phenotype with anticipated phenotypes in a CP patient population. In some cases, a wide clinical spectrum is associated with the gene in the literature and the anticipated patient population. In this case, the middle of the spectrum was used to score severity.

**Effect size, bias assessment, and protocol registration**

Effect size was not considered since the types of data reported in individual studies did not allow for meta-analyses. We did not systematically assess the possibility of bias. We coud not control for the higher likelihood of the publication of positive findings (i.e. effective treatments) over negative findings (non-effective treatments). The nature of the literature search also means we likely missed reports that were not linked to the gene name and search terms used in Pubmed, which likely results in a more conservative report where actionable findings were overlooked.

This study was not registered and a protocol was not prepared or reviewed in advance.

**Additional web resources**

Oxford Centre for Evidence-Based Medicine guidelines

https://www.cebm.ox.ac.uk/resources/levels-of-evidence/ocebm-levels-of-evidence

CTCAE

https://ctep.cancer.gov/protocoldevelopment/electronic_applications/docs/ctcae_v5_quick_reference_5x7.pdf

MTBQ

https://www.bristol.ac.uk/primaryhealthcare/resources/mtbq/

**Supplemental Data**

**Supplemental Figure 1** Criteria for assigning actionable gene finding categories, design of clinical utility impact rubric, and scoring process.

**Supplemental Table 1** list of definitions used in the study

**Supplemental Table 2** List of 243 genes identified in individuals with CP undergoing ES

**Supplemental Table 3** Inter-rater reliability

**Supplemental Excel** Outcome-intervention pairs with gene name, OMIM phenotype number, assigned scores, references used for evidence, and categories mechanisms of intervention.

### A Category assignment process

| Primary | Preventative | Symptom | Future |
|---|---|---|---|
| target disease mechanism (ex. replacing biochemical deficiency, gene therapy) | avoid triggers that worsen function or require surveillance | identify an effective treatment w/ reduced trial and error (ex. candidates for DBS or ketogenic diet) | pharmaceuticals in development (clinical trial, druggable genome) |

Literature-based assignment process:
1. Genes assigned to single category based on highest ranking (primary>preventative> symptom> future)
2. Mechanism assigned based on published molecular target of the intervention and effect on the outcome
3. Clinical trials include Phase II/III either in process, concluded, or with the compound undergoing FDA review. Terminated trials excluded.

### B Impact assessment process

| Evidence | Severity | Nature | Efficacy |
|---|---|---|---|
| quality of literature/extent of study of intervention | impact of outcome on patient function (80 unique outcomes) | burden or risk of intervention (81 unique interventions) | magnitude of improvement of intervention on outcome |

Modified Delphi process:
1. Formed working group of genetic counselors, neurologists, developmental pediatricians, and clinical and research geneticists.
2. Built rubrics (from ClinGen framework)
3. Virtual discussion and written rubric revisions/feedback
4. Iterative individual scoring
5. Virtual discussion and scoring consensus where needed

**Supplemental Figure 1** Criteria for assigning actionable gene finding categories, design of clinical utility impact rubric, and scoring process. **A**, Process for assigning categories for type of intervention. Interventions were assigned to a single category based on the highest rank (with primary being higher ranked than prevention, and finally symptom management). Interventions from the future classification were not assessed for clinical impact. **B**, Process for developing rubric and strategies for evaluating clinical impact of precision medicine finding.

| Term | Definition |
|---|---|
| Outcome | Potential or ongoing clinical concern affecting patient |
| Intervention | A personalized medicine approach that becomes applicable after the identification of the genetic etiology (not part of the standard of care) |
| Severity | How the condition will impact patient function if untreated |
| Risk | Potential risk of medical or surgical treatment, such as the severity of a potential adverse event (AE) |
| Burden | The impact to patient/family, such as invasiveness, significant lifestyle alterations, or requiring specialized management |
| Efficacy | The degree to which an intervention can prevent or significantly reducing symptoms, improving function, or decrease risk of additional complications/manifestations |

**Supplemental Table 1** Definitions used in study

| | | | | | |
|---|---|---|---|---|---|
| *ABCC8* | *CIT* | *FUCA1* | *MED13L* | *REEP1* | *TBL1XR1* |
| *ACTB* | *CLTC* | *G6PD* | *MEF2C* | *RNASEH2B* | *TCF4* |
| *ACTG1* | *CLTCL1* | *GABRA1* | *MFN2* | *RPE65* | *TELO2* |
| *ADAR* | *COL4A1* | *GABRB2* | *MICU1* | *RYR1* | *TOE1* |
| *ADCY5* | *COL4A2* | *GFAP* | *MOCS2* | *RYR2* | *TPP1* |
| *ADNP* | *CREBBP* | *GLRA1* | *MTFMT* | *SACS* | *TRAPPC9* |
| *AFG3L2* | *CTBP1* | *GNAO1* | *NAA10* | *SATB2* | *TREX1* |
| *AHDC1* | *CTC1* | *GNAS* | *NACC1* | *SCN1A* | *TRIP12* |
| *AIFM1* | *CTNNB1* | *GNB1* | *NALCN* | *SCN2A* | *TSEN54* |
| *ALDH3A2* | *CUL4B* | *GPT2* | *NGLY1* | *SCN8A* | *TUBA1A* |
| *AMPD2* | *CYP2U1* | *GRIA3* | *NIPBL* | *SEPSECS* | *TUBB2A* |
| *AP1S2* | *DDC* | *GRIN1* | *NKX2-1* | *SETBP1* | *TUBB2B* |
| *AP4M1* | *DDHD2* | *GRIN2B* | *NKX6-2* | *SETD2* | *TUBB3* |
| *AP4S1* | *DDX3X* | *HACE1* | *OPA1* | *SETD5* | *TUBB4A* |
| *ARID1B* | *DKC1* | *HDAC8* | *OPHN1* | *SHANK3* | *UBA5* |
| *ARID2* | *DNM1* | *HECW2* | *OTUD6B* | *SHOC2* | *UBE3A* |
| *ARSE* | *DNM1L* | *HIST1H1E* | *PANK2* | *SIL1* | *UPF3B* |
| *ARX* | *DNMT3A* | *HNRNPH2* | *PCDH12* | *SIN3A* | *USP9X* |
| *ASXL1* | *DOCK6* | *HPRT1* | *PDHA1* | *SLC13A5* | *VPS11* |
| *ASXL3* | *DYNC1H1* | *IDS* | *PDHX* | *SLC16A2* | *WAC* |
| *ATL1* | *DYRK1A* | *IFIH1* | *PEX16* | *SLC1A2* | *WDR26* |
| *ATM* | *EARS2* | *IQSEC2* | *PGAP1* | *SLC1A4* | *WDR45* |
| *ATP1A2* | *EBF3* | *ITPR1* | *PHIP* | *SLC2A1* | *WWOX* |
| *ATP1A3* | *ECHS1* | *KAT6A* | *PHKA1* | *SLC6A3* | *YY1* |
| *ATP5A1* | *EEF1A2* | *KCNB1* | *PIEZO2* | *SMARCA2* | *ZC4H2* |
| *ATP6V0A4* | *EHMT1* | *KCNC3* | *PIGA* | *SMARCB1* | *ZDHHC9* |
| *ATP7A* | *EIF2B2* | *KCNQ2* | *PIGN* | *SNAP29* | *ZEB2* |
| *ATRX* | *EP300* | *KCNT1* | *PIGV* | *SON* | *ZNF423* |
| *AUTS2* | *EPG5* | *KCTD7* | *PIK3CD* | *SOX2* | |
| *BCAP31* | *ERCC6* | *KDM5C* | *PLA2G6* | *SPAST* | |
| *BCL11A* | *ERCC8* | *KIAA2022/NEXMIF* | *PLOD1* | *SPATA5* | |
| *C5orf42* | *EXOSC3* | *KIDINS220* | *PLP1* | *SPG11* | |
| *CACNA1A* | *EZH2* | *KIF11* | *PNKP* | *SPR* | |
| *CACNA1G* | *FA2H* | *KIF1A* | *POGZ* | *SPTAN1* | |
| *CALM1* | *FAR1* | *KMT2A* | *POLG1* | *STAMBP* | |
| *CAMTA1* | *FARS2* | *KMT2B* | *POMGNT1* | *STAT3* | |
| *CASK* | *FBN2* | *L1CAM* | *PPM1D* | *STXBP1* | |
| *CDC42* | *FGD1* | *LIPH* | *PPT1* | *SUMF1* | |
| *CDKL5* | *FGF12* | *LMAN2L* | *PRUNE1* | *SYNE1* | |
| *CHD2* | *FGFR1* | *MAP2K1* | *PTPN11* | *SYNGAP1* | |
| *CHD8* | *FH* | *MBD5* | *PURA* | *TAF1* | |
| *CHMP1A* | *FOXG1* | *MECP2* | *RAB3GAP1* | *TANGO2* | |
| *CHRNE* | *FRRS1L* | *MED12* | *RAB3GAP2* | *TBCK* | |

**Supplemental Table 2**: List of 243 genes with P/LP variants in 1841 individuals sequenced in either clinical or research setting included in the analysis.

| metric | # scorers | # outcomes/ interventions | percent agreement | AC2 coefficient | p value |
|---|---|---|---|---|---|
| severity | 3 | 16 | 94% | 0.89 | $1.43x10^{-07}$ |
| | 4 | 21 | 87% | 0.74 | $1.41x10^{-10}$ |
| | 5 | 4 | 90% | 0.81 | $1.68x10^{-04}$ |
| | 6 | 4 | 86% | 0.71 | $4.46x10^{-04}$ |
| | 7 | 5 | 80% | 0.56 | $3.29x10^{-03}$ |
| | 8 | 12 | 78% | 0.47 | $6.97x10^{-06}$ |
| | 9 | 9 | 78% | 0.53 | $1.78x10^{-05}$ |
| risk/ burden | 3 | 4 | 100% | 1 | 0 |
| | 4 | 20 | 90% | 0.81 | $3.10x10^{-11}$ |
| | 5 | 33 | 92% | 0.81 | 0 |
| | 6 | 5 | 78% | 0.41 | $1.80x10^{-02}$ |
| efficacy | 3 | 70 | 93% | 0.83 | 0 |
| | 4 | 20 | 85% | 0.63 | $3.87x10^{-09}$ |
| | 5 | 9 | 85% | 0.61 | $6.65x10^{-04}$ |
| | 6 | 2 | 64% | 0.16 | 0.2 |
| Average | | | 85.5% | 0.66 | |
| CI | | | 81.0-89.6% | 0.56-0.77 | |

**Supplemental Table 3**: Interrater reliability. The analysis was performed based on the number of scorers who contributed to the final average for each outcome/intervention. The Gwet AC2 statistic ranges from -1 (complete disagreement) to +1 (complete agreement). The coefficient is interpreted based on the range described for Landis and Koch's interpretation for Kappa, where 0.41-0.6 is moderate agreement and 0.61-0.80 indicates substantial agreement. Only one comparison did not reach statistical significance ($p<0.05$), which were 2 efficacy outcome-interventions ratings that required 6 scorers and a consensus discussion to finally reach consensus. Linear regression did not identify any relationship between the # of scorers or # of outcome-intervention pairs on percent agreement, the AC2 coefficient, or the p-value (data not shown).
